# The Risk of Mpox (Monkeypox) Importation and Subsequent Outbreak Potential in Mainland China: A Retrospective Statistical Modelling Study

**DOI:** 10.1101/2023.08.24.23294530

**Authors:** Xiaowei Deng, Yuyang Tian, Junyi Zou, Juan Yang, Kaiyuan Sun, Hongjie Yu

## Abstract

The 2022 mpox outbreak has spread rapidly across multiple countries in the non-endemic region, mainly among men who have sex with men (MSM), while China only has limited recorded importation and no local outbreak. We constructed probabilistic models to simulate the risk of mpox importation in mainland China, with the help of reported monkeypox cases during this multi-country outbreak and the international air-travel data. And we further evaluated the mpox outbreak potential given that undetected mpox infections were introduced into men who have sex with men, considering different transmissibility, population immunity and population activity. We found that the reduced international air-travel volume and stringent border entry policy decreased about 94% and 69% mpox importations respectively. Once a mpox case is introduced into active MSM population with almost no population immunity, the risk of triggering local transmission is estimated at 42%, and would rise to >95% with over six cases. Our study demonstrates the key role of the reduced international air-travel volume and stringent border entry policy during the COVID-19 pandemic on reducing mpox importations, and the subsequent risk of triggering local outbreaks among MSM.

## Introduction

Since the first human infection was detected in Central Africa in 1970(*1*), the monkeypox virus has been recognized as a viral zoonosis with endemic regions primarily concentrated in west and central African countries(*2, 3*). The monkeypox virus, a member of the *Orthopoxvirus* genus in the Poxviridae family, is closely related to the widely known variola virus (which causes smallpox)(*4*). In the last two decades, the global incidence of monkeypox cases (here after, monkeypox is denoted as mpox according to WHO reference) has been on the rise in endemic regions, with an increase in the median age among confirmed and probable mpox cases over time(*5*). Before the 2022 multi-country mpox outbreak, imported mpox infections in humans have been reported in the USA(*6-9*), UK(*10*), Israel(*11*), and Singapore(*12*).

After the confirmation of a mpox case in England on May 7, 2022, the outbreak has rapidly expanded spatially to other countries in Europe, the Americas, and Western Pacific region(*13*). It is the first time that mpox cases have been reported in both endemic and non-endemic regions with broad geographical coverage. This multi-country mpox outbreak also reveals different epidemiological and clinical characters from previous outbreaks(*7, 14-18*) (Table S1). In particular, sustained human-to-human transmission has been observed, involving predominately men who have sex with men (MSM)(*19*), which made mpox as a sexually transmitted disease (STD)(*20*). On July 23, 2022, the WHO declared the multi-country outbreak of mpox as a Public Health Emergency of International Concern (PHEIC) due to its unprecedent rapid spread in the non-endemic regions and disparate global dissemination(*21, 22*), and since May 11, 2023, the WHO cancel the PHEIC declaration. Recently, countries/regions in East Asia has started experiencing local mpox outbreaks that were not seen during the height of mpox outbreak in 2022(*23*). As of June 15, 2023, 181 confirmed cases have reported by Japan and 102 cases by the Republic of Korea as of June 5(*24, 25*). Taiwan province reported over 50 local mpox cases in May 2023. It is crucial to evaluate the mpox outbreak potential in mainland China, due to its large pool of susceptible population at risk of mpox and the fundamental change in the geographical range of mpox circulation.

In this study, we used mathematical models to evaluate: 1) the importation risk by international air travel from countries with ongoing outbreaks; 2) the effectiveness of two sets of border screening strategies considering mainland China’s pre- and peri-pandemic (hypothetical) border entry and quarantine policies; 3) the risk of undetected mpox importations to trigger a local outbreak in mainland China across a range of plausible mpox transmissibility and contact patterns among the MSM community.

## Results

According to the WHO situation report(*26*), as of June 13, 2023, a total of 87,979 confirmed mpox cases were reported among 111 countries and regions, with the majority of cases reported in the American (59,449) and European (25,910) regions, accounting for 67.6% and 29.5% of the global cases (Fig.1).

**Fig. 1.**
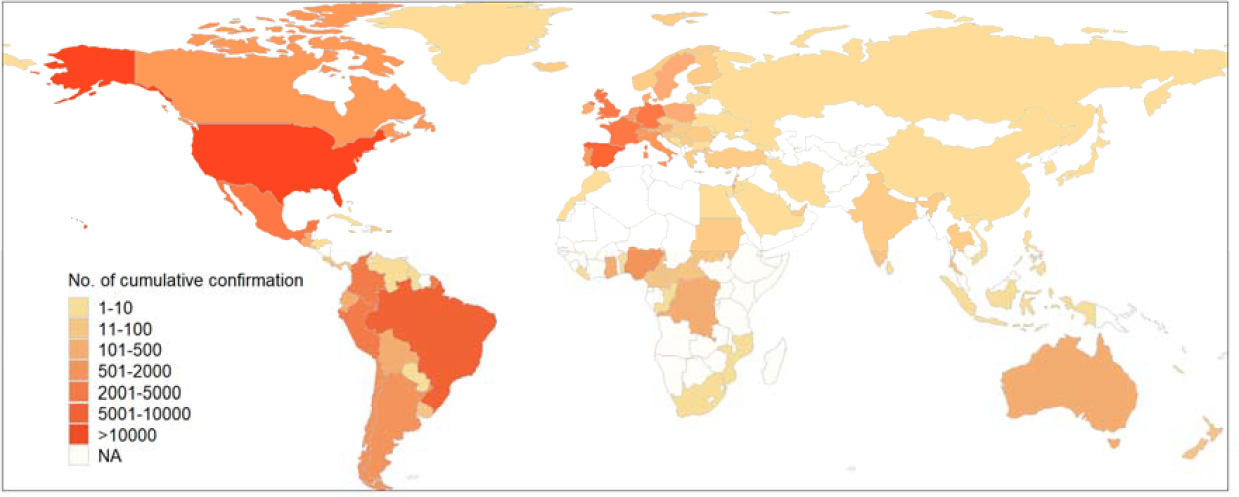
Spatial distribution of mpox cases. Spatial distribution of confirmed mpox cases from January 1, 2022 to June 13, 2023. Shapes in color of ivory indicate that data in countries/regions is not available (NA).

According to the air-travel data from OAG(*27*), we found the monthly number of arrivals in China was on average 5.69 million during 2017–2019. However, the volume drastically decreased to 0.6 million starting in January 2020, approximately one-tenth of the pre-pandemic (2017–2019) air-travel volume (Fig.2a, inset). Assuming a hypothetical scenario that the year 2022 has the same air-travel volume as in 2019 (pre-pandemic), we estimated that the expected number of mainland China’s mpox importations would have risen above one on June 2, 2022 and reached 36 (95% CI: 27–50) on September 11, 2022. In contrast, according to the actual international air travel volume during 2022 (peri-pandemic), we estimated that the expected cumulative importations would only reach two (95% CI: 0–5) by September 11, 2022. In Fig.2b, we presented mainland China’s top 12 source countries for mpox importations based on the travel volume of year 2019 and 2022, respectively. Under the air-travel volume of 2019, mainland China would expect (on average) more than one mpox infection importation from epidemic countries including the United States (16, 95% CI: 8–23), the United Kingdom (7, 95% CI: 3–12), Spain (4, 95% CI: 1–8), France (4, 95% CI: 0–8), Canada (2, 95% CI: 0–5), Germany (1, 95% CI: 0–4) and Netherlands (1, 95% CI: 0–3). In contrast, under the air-travel volume of 2022, we estimated that no single source country would contribute to more than one mpox importation to mainland China, with the highest-ranking country (the United States) contributing to about one mpox importation (95% CI: 0–3).

**Fig. 2.**
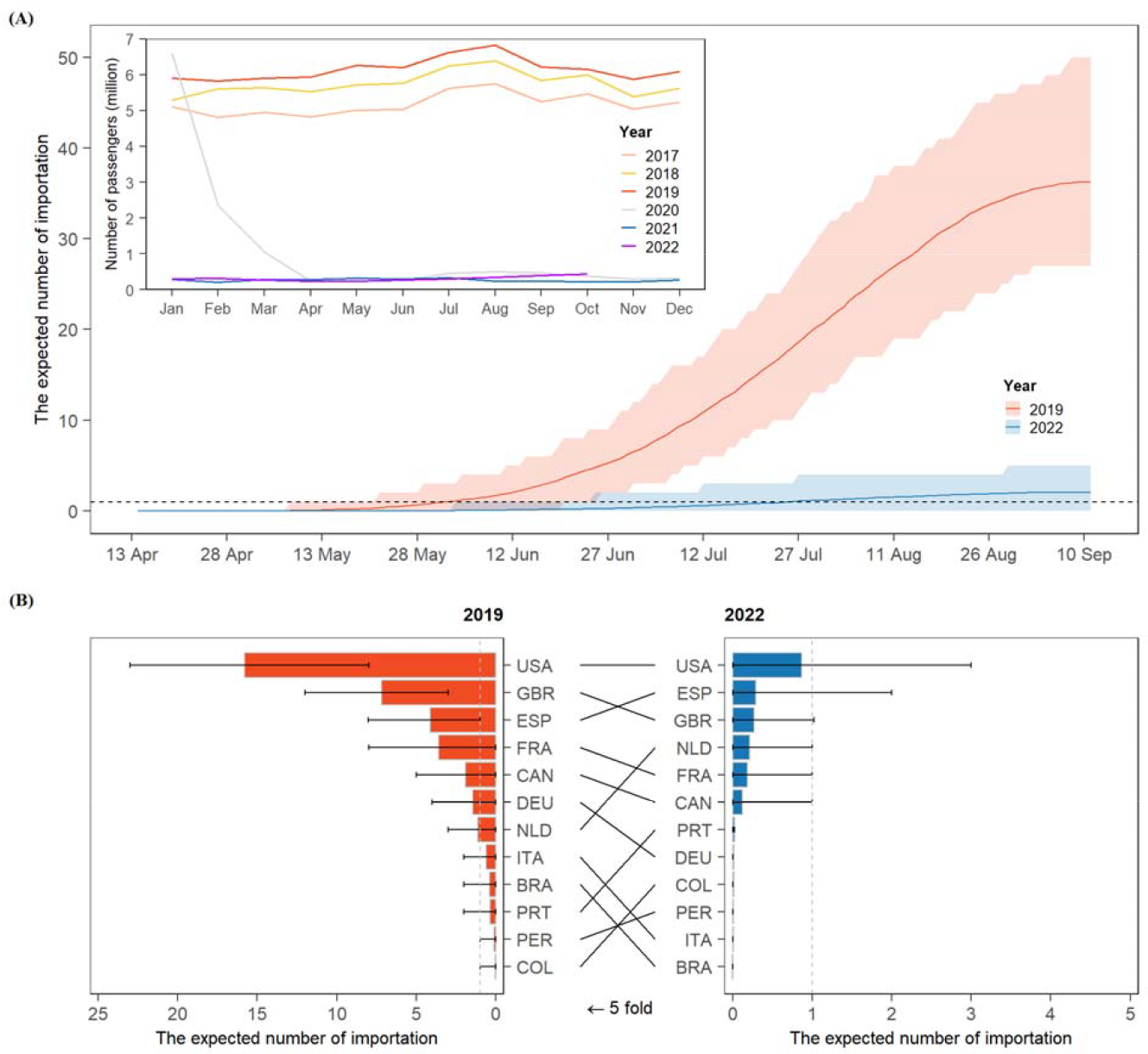
The air-travel volume and the expected number of mpox importations in China. (A) The expected cumulative number of mpox importations from May to September 2022 with air-traffic volume in 2019 and 2022. (B) The expected number of mpox case importation from other countries/regions with air-traffic volume in 2019 and 2022. In panel a, the inset indicates the monthly air-travel volume to mainland China from 2017 to 2022. In panel b, the three letter texts are ISO Alpha-3 codes, i.e., DEU denotes Germany, ESP denotes Spain, GBR denotes the UK, etc. Detailed ISO Aplha-3 codes can be seen in Table S8.

We first examined the effectiveness of a mpox border screening policy (measured as the probability of mpox infection detection among all mpox importations) under mainland China’s pandemic quarantine policy for international travelers (Scenario 1). Specifically, we found that under Scenario 1, the effectiveness of border screening against mpox is insensitive to the self-reporting rate of a mpox epidemiological links, as all international travelers were required to quarantine due to existing pandemic policies. On the other hand, the detection rate of mpox importations increases with the quarantine duration (Fig.3a). Mainland China’s pandemic policy requiring a quarantine duration of 10 days (policy as of June 28, 2022(*28*)) would lead to high effectiveness of mpox screening with a mpox detection rate reaching >80% based on our model’s estimate, irrespective of the self-reporting rate of epidemiological links. However, universal border entry quarantine as a stand-alone policy for mpox, in the absence of pandemic-related policy, is impractical and unwarranted due to mpox’s much slower spread compared to COVID-19 and the lack of evidence of pre-symptomatic transmission(*29*). We thus considered a scenario of a border screening policy similar to the contact tracing guidance by the European Center for Disease Prevention and Control (ECDC)(*29*). In this scenario, both the self-reporting rate of epidemiological links and the duration of the medical observation influence the screening effectiveness: the proportion of imported mpox infections who self-report an epidemiological links sets the upper bound of the mpox border screening effectiveness, as infected individuals who do not report/are unaware of mpox exposure would not undergo medical observation and thus could not be promptly detected (Fig.3b). Among exposed individuals undergoing medical observation, increasing the duration of medical observation would increase the detection rate of border screening but the increase would start plateauing with only marginal benefit once the duration reached beyond 10 days. In Fig.3c, we compared the overall effectiveness of Scenario 1 against Scenario 2 across a range of quarantine/medical observation durations, conditional on the same proportion of self-reporting epidemiological link (50%). We found that the more stringent Scenario 1 always outperforms Scenario 2 across different quarantine/medical observation durations. At the quarantine/medical observation duration of 10 days, Scenario 1 on average detects 83.3% of imported mpox cases while Scenario 2 could only detect 46.3%. Longer quarantine/medical observation duration does not significantly improve the border screening effectiveness for either of the two scenarios.

**Fig. 3.**
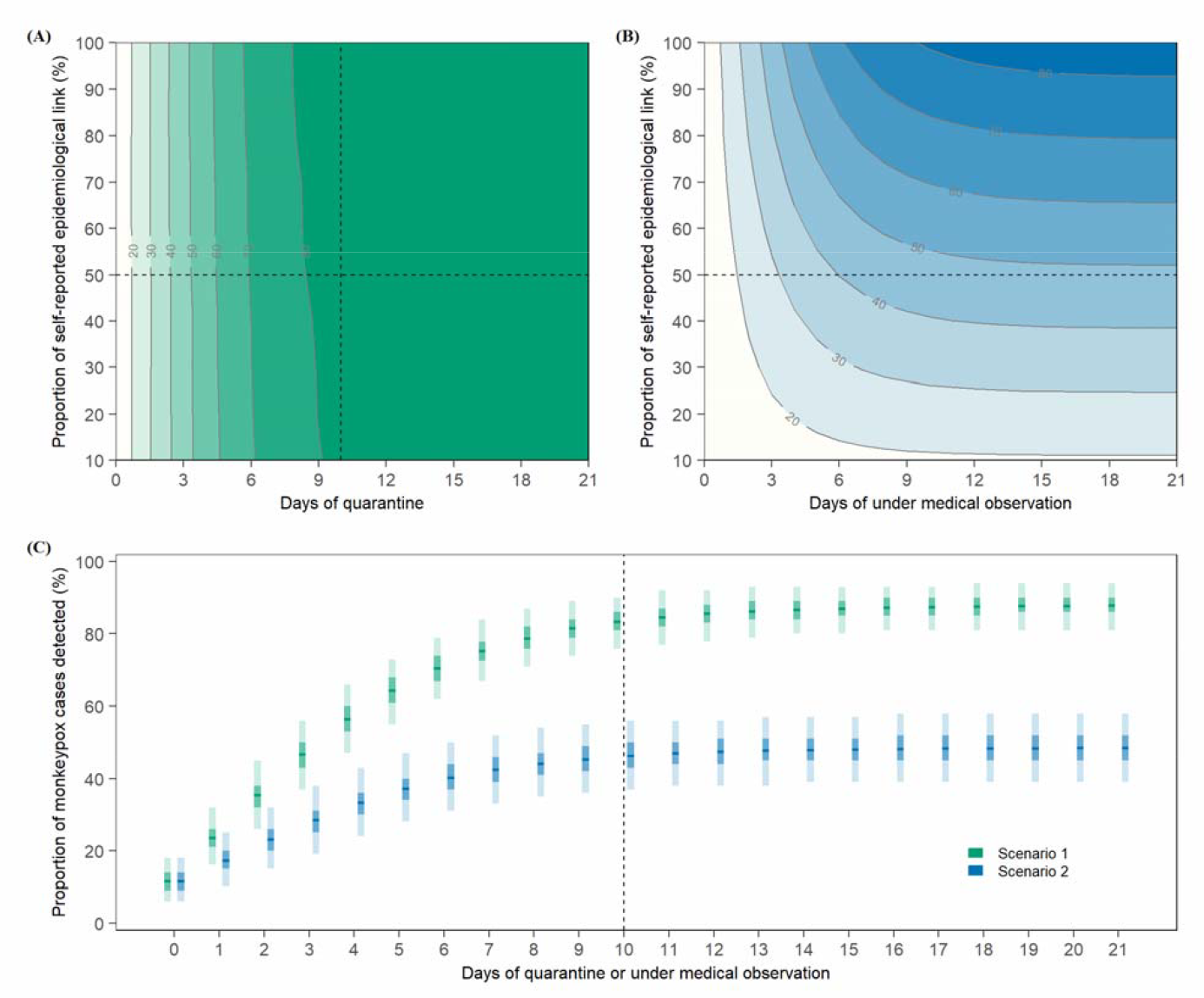
Effectiveness of mpox border screening policies under two assumed scenarios. (A) Number of total detected mpox cases under Scenario 1 (corresponding to China’s pandemic quarantine policy), with proportion of self-reported epidemiological links ranging from 10% to 100%, and days of quarantine ranging from 0 to 21. (B) Number of total detected mpox cases under Scenario 2 (a pre-pandemic scenario), with proportion of self-reported epidemiological links ranging from 10% to 100%, and days of medical observation ranging from 0 to 21. (C) Number of total detected mpox cases with 50% self-reported epidemiological links with mpox cases under days of quarantine or medical observation from 0 to 21. In panel c, the horizonal lines indicate the mean, with dark and light shades indicating 50% CI, and 95% confidence interval.

Next, we used a mathematical model to evaluate the potential of undetected mpox infections triggering a local mpox outbreak. The outbreak potential of an undetected mpox importation is influenced by the overall transmissibility of the mpox virus (measured as the basic reproduction number *R*_0_) and the transmission heterogeneity (measured as the overdispersion parameter *k*, Supplement, section 8)(*30*). In Fig.4a, we demonstrated the outbreak probability of a single undetected mpox infection in the high-risk population as a function of *R*_0_ and *k*: both the increasing of *R*_0_ (increasing transmissibility) and *k* (decreasing transmission heterogeneity) would increase the outbreak probability. Considering the recent estimation of *R*_0_=1.8 for mpox outbreaks in several European countries, the outbreak probability caused by one undetected mpox case was estimated at 42.1% if the overdispersion parameter *k* was equal to 0.88, based on sexual contact network among MSM in mainland China(*31, 32*). The outbreak probability drastically goes up with the number of undetected mpox importations, reaching to above 95% with six or more importations. When taking into account of emergency vaccination, the reproduction number is reduced to 1.46 and the outbreak probability accordingly reduces to 29.4% minimum, which is 70% of the situation with *R*_0_=1.8.

**Fig. 4.**
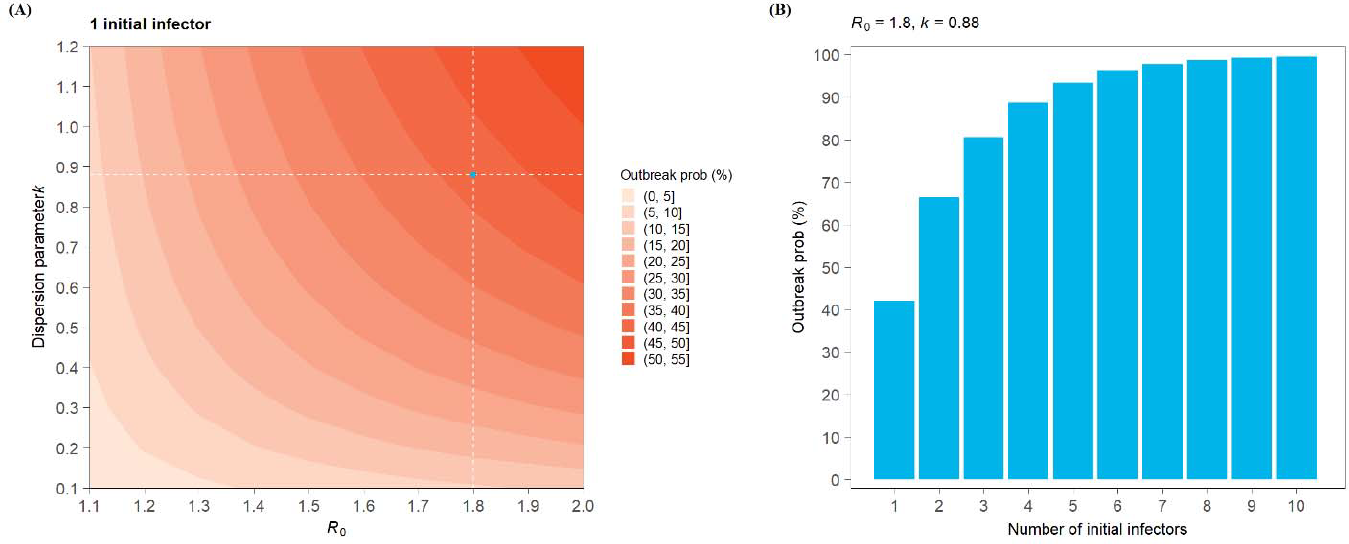
Estimated outbreak probability. (a) the outbreak probability of one mpox case introduced into the MSM community in mainland China, with R0 ranges from 1.1 to 2, and dispersion parameter k ranges from 0.1 to 1.2. In panel a, the dashed lines and dot refer to *R*_0_=1.8 and k=0.88, a plausible guess of mpox transmissibility in mainland China. (b) the outbreak probability as a function of the number of seeding infectors ranges from 1 to 10, with *R*_0_=1.8 and k=0.88.

## Discussion

Since the global eradication of smallpox in the 1980s and subsequent halt of the smallpox vaccination program, it has been suggested that the mpox virus could emerge as the next *Orthopoxvirus* of epidemic potential in humans, taking up the ecological niche vacated by smallpox in the human population(*33*). In recent years, increasing incidence and shifting demographics towards older populations and males in the endemic region were warning signs of declining of mpox population immunity due to the cessation of smallpox vaccination in the 1980s and population turnover. Following the first confirmed mpox case reported by the United Kingdom in early May 2022, the multi-country mpox outbreak had infected about 86,000 individuals in non-endemic regions as of June 13, 2023, with countries in Europe and the Americas being impacted most heavily. After confirming mpox’s local transmission, the UK and other European countries subsequently adopted contact tracing and case isolation to control their outbreaks(*34, 35*). Following the PHEIC declaration of the outbreak by the WHO, the US government declared a public health emergency for the local mpox outbreak and adopted measures (case identification, contact tracing, vaccination) to suppress local transmission. When faced with the importation risk of mpox, only a few countries have strengthened their border screening to prevent case importation(*36-41*).

On September 16, 2022, mainland China recorded its first imported mpox case in Chongqing. The case had a travel history to Germany and Spain(*42*) and remained the only observed mpox importation as of June 5, 2023. This observed low risk of mpox importation is in line with our model’s projections based on 2022 air-travel volume. By examining the OAG’s data through October 2022, mainland China’s air travel has yet to show significant recovery from that of 2022 (Fig.2a) due to the stringent border entry policy that is still in place. Furthermore, as a byproduct of the universal quarantine policy for border entry due to the pandemic, our analysis suggests high border screening effectiveness for mpox if medical observation for mpox were implemented during the mandatory quarantine. Collectively, the impact of pandemic on air-travel reduction and a stringent quarantine policy have led to a low risk of mpox importation in mainland China from the current multi-country mpox outbreak. However, under conditions where air-travel volume is able to rebound to pre-pandemic levels, we estimate that the importation risk is substantially higher, reaching 36 (95% CI: 25–47) mpox importations under the current outbreak (Fig.2a).

A distinctive feature of the 2022 mpox outbreak is that it affects predominantly the population in the MSM community with sustained human-to-human transmission outside the endemic region(*13*). Modeling analysis of the UK’s sexual contact network of the MSM community suggested the heavy-tailed nature of network topology could result in the basic reproduction number of mpox larger than the epidemic threshold of one among the group of individuals who have disproportionately large numbers of partners(*43*). We have looked at the distribution of sexual partners among MSM in China in 2010 based on the study by Zhang et. al.(*31, 32*) (Fig.S3); it is highly heterogeneous, with 4.9% of individuals having more than 10 sexual partners over a period of one year. Thus, it is plausible to expect that once mpox infection is introduced into the MSM community in mainland China, the mpox virus could sustain human-to-human transmission and cause substantial disease burden in the high-risk population. Assuming the mpox’s basic reproduction number in mainland China is similar to that in Europe and the United States(*44*), we estimate that the probability of triggering a local mpox outbreak is substantial and rapidly goes up with the number of mpox introductions (Fig.4). A recent study suggests that the size of the MSM community in mainland China is approximately eight million(*45*), roughly twice the size of that in the US(*46*). With the changing epidemiology and population immunity of mpox as well as the geographical expansion to non-endemic regions, mpox has emerged as a public health threat for mainland China’s MSM community in the foreseeable future.

Although the global mpox epidemic trend has been on the decline since August, 2022, and the WHO declared that the mpox epidemic no longer constituted a PHEIC, mpox is still circulating with low intensity in non-endemic countries/regions recently(*22*). Several Asian countries including Japan, Thailand, etc. have reported imported and local mpox cases since May, 2023(*23, 47*). Taiwan Province reported over 50 local mpox cases in May, 2023. From June to September, 2023, 29 provinces in mainland China reported 1,043 confirmed mpox cases, suggesting likely wide-spread mpox local outbreaks in mainland China. This is in agreement with our modeling assessment of high mpox outbreak probability in mainland China, conditional on introduction of travel-related mpox cases (Fig.2). We explored the correlation between the confirmed mpox cases and active MSM population size at the province level estimated by Hu et al(*45*), if we remove Beijing and Guangdong province (both are major international air traffic hubs for mainland China) presented as outliers (Fig.S4). We further performed a regression model including both mpox cases active MSM population size and air-travel volume as covariate (with interaction), without removing Bejing and Guangdong (Supplement, section 10). The result of the regression showed high coefficient of determination (R^2^ = 0.8180), indicating that both the active MSM population size and air-travel volume contribute to explaining mpox cases number.

Our study has several limitations. First, we assumed that the air-travel behavior of the population in the MSM community is the same as that of the general population, which may not be true. Second, we assumed there is no under-reporting of mpox infections, which may lead to an underestimation of the importation risk in mainland China. However, considering the high symptomatic rate of mpox infection as well as distinct symptom presentations including fever and rash, we expect our estimation and our conclusion remain robust. Lastly, data on MSM sexual contact networks are limited in mainland China. In particular our MSM sexual contact network is based on data pre-pandemic (2010), and during the pandemic, the sexual behavioral could have changed substantially. Better characterization of the behavioral aspect of the high-risk population could further improve our evaluation of mpox outbreak potential and define effective preventive and mitigation public health policies against mpox.

Using pre- and peri-pandemic air-travel data and spatiotemporally resolved mpox case reporting, our study demonstrates that the reduced international air-travel volume and stringent border entry policy during the COVID-19 pandemic reduced mpox importations prominently. However, the risk could be substantially higher with the recovery of air-travel volume to pre-pandemic level. Mpox could emerged as a public health threat for mainland China given its large MSM community.

## Materials and Methods

### Simulating the international dissemination of the 2022 multi-country mpox outbreak and evaluating mainland China’s importation risk

To quantify the international importation risk of mpox from a source location *i* (regions with ongoing mpox transmission) to a destination location *j*, we need to know the outbreak size in the source location *i* and the connectivity between the origin (location *i*) and destination (location *j*) by air-travel. We assumed that travel-associated mpox cases that contribute to the international spread mostly reside in major metropolitan areas with international airport transportation hubs. Hence, we considered a total of 39,114 mpox cases which occurred in 39 metropolitan areas with major international airports in the top 12 countries affected by mpox (Tables S2 and S3) and evaluate their risk of exportations to mainland China. Specifically, we obtained the time series of mpox incidence at the national level from Global.health(*48*). We then decomposed the national-level time series into metropolitan-level proportional to the relative outbreak size of a metropolitan area with respect to its national total. For each mpox infection at the source location *i*, we assume that the infected individual could only make international air travel during the incubation period of τ days (i.e., prior to symptom onset), which we drew from a lognormal distribution with an average of 8.94 days and standard deviation of 4.19 (Table S4). For each case, we assumed there is a delay of σ days between symptom onset and the date of reporting, drawing from a zero-inflated negative binomial distribution with an average of 6.48 days and a standard deviation of 28.2 days (Supplement, section 4, Fig.S1). Consequently, if a mpox case is reported on date, this infected individual could potentially travel internationally between date *t* − σ and date *t* − σ − *τ*, i.e., during his/her incubation period.

We then estimated the international air travel probability using the origin-destination air-travel data provided by Official Aviation Guide (OAG). The pandemic greatly influenced global air travel. We considered a pre-pandemic (upper-bound) scenario using the air-travel data of year 2019 and a peri-pandemic (lower-bound) scenario using the air-travel data of year 2022. For each air-travel scenario, if we denote the total air-travel volume at month *m* from location *i* to *j* location as 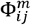 (provided by OAG), the per-capita rate of international air travel Φ_*ij*_ can be expressed as 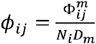, where *N*_*i*_ is the total population in location *i* and *D*_*m*_ is the number of days in month *m*. We then iterated through all mpox cases and simulated each of their potential air-travel trajectory during their incubation period τ: starting from the beginning of the incubation period *t* − σ − τ, for each day forward *t*′ till the end of the incubation period *t* − σ, whether the individual would travel on date *t*′ is simulated as a Bernoulli process with the probability of travel *p*_*i*_(*t*′)from location *i* is given by 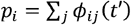. Given that the individual would travel on *t*′, then the destination *j* of the international travel would be decided based on a generalized Bernoulli distribution with the conditional probability of traveling to a specific location *j* given by 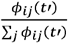 and the simulation for this infected individual ends. The simulation of the infected individual would also end if by the end of his/her incubation period *t* − σ, the infected individual does not travel, and this individual would remain as a local mpox case in location *i*. Once we iterate through all mpox infections globally, we could sum up the cumulative number of mpox importations from location *i* to location *j* up to time point *t*, denoted as *m*_*ij*_ (*t*). We repeated the entire simulation for a total of 200 times and obtained the distribution of *m*_*ij*_ (*t*), denoted as *p* (*m*_*ij*_ (*t*)) to capture the stochastic nature of the modelled process. We could estimate the mean and 95% confidence interval (0.025 and 0.975 quantiles) of *m*_*ij*_ (*t*) based on *p* (*m*_*ij*_ (*t*)). To ensure the validity of our model, we performed a validation analysis of the mpox importations among countries where consecutive reports of mpox cases with international travel history were available, with abovementioned pre-pandemic and peri-pandemic air-travel volume setting the upper bound and lower bound of the importation risks, respectively (Fig.S2).

### Effectiveness of border screening against travel-associated mpox cases

To quantify the effectiveness of border screening policies, we first considered a scenario (denoted as Scenario 1 hereafter) that is in compliance with mainland China’s current pandemic quarantine policy for border entry: according to the recently published (July 1, 2022) guidelines to mpox prevention and control by mainland China’s National Health Commission, all international travelers entering mainland China, especially for those who had travel history to regions with an on-going mpox epidemic within 21 days of border entry, should be screened for mpox during their mandatory quarantine as a part of mainland China’s pandemic response(*49*). During the quarantine, individuals are also screened for mpox related symptoms including fever and rash. Symptomatic individuals with travel history from outbreak countries, close contact with confirmed cases, or contact with infected animals would be defined as suspected cases (Table S5). Once detected, they would be reported to the local CDC and will be transferred to designated hospitals for a 21-day medical observation, and PCR test for lesion samples/swabs/blood will be conducted. Considering the variation of pandemic mandatory quarantine durations (ranging from 7–21 days) as well as the uncertainty on the proportion of people with mpox infections complying with self-reporting, we used a mathematical model to evaluate the proportion of imported mpox infections detected post entry screening (Supplement, section 7). We also considered another hypothetical scenario (denoted as Scenario 2 hereafter) of plausible mpox entry screening in the absence of mainland China’s pandemic quarantine policy for international travelers (a pre-pandemic scenario). In this scenario, international travelers with a self-reported mpox epidemiological link needed to undergo medical observation for mpox-related symptoms (e.g., fever, rash, lymphadenopathy) during which they abstain from sexual activity, in addition to other precautionary measures(*29*). Upon symptom onset, a suspected mpox case needs to self-isolate and seek testing. We also evaluated the effectiveness of this travel screening policy (Supplement, section 7).

### Local outbreak probability

Following Hartfield et al.’s analytical solution of outbreak probability as a function of the basic reproduction number *R*_0_ and overdispersion parameter *k*(*30*) (Supplement, section 8), we explored the range of mpox outbreak probability with one initial infector introduced into the high-risk population in mainland China, with *R*_0_ ranging from 1.1 to 2.0 and dispersion parameter *k* ranging from 0.1 to 1.2 (Fig.4a). Our best guess for *R*_0_ for the 2022 mpox outbreak is 1.8, according to a recent study conducted by Kwok et al(*44*), and we consider a plausible dispersion parameter *k* of 0.88 through fitting the negative binomial distribution for the number of sexual partners among MSM in China(*31, 32*) (Fig.S3). For *R*_0_ =1.8, and *k = 0*.*88*, we explored how the outbreak probability would change with the number of imported mpox cases missed by the border screening process (Fig.4b). To understand the effect of emergency vaccination on preventing the outbreak probability, we used *R*_t_ to stand for the protection provided by emergency vaccination. The relationship can be illustrated as *R*_0_ = *R*_0_ * *S*, in which *S* stands for the proportion of population susceptible to mpox. According to the US emergency vaccination of mpox as well as the vaccine effectiveness(*50, 51*), we assumed 20% high risk population get effectively vaccinated, hence the *R*_0_ =1.46.

All analysis was done in R (version 4.1.1).

## Data Availability

The data that support the findings of this study are available from the corresponding author, upon reasonable request.

## Acknowledgments

We would like to thank our team members (Yan Wang, Lan Yi, Jiayi Dong, Qianhui Wu, and Jun Cai) for communications on this study. The findings and conclusions in this report are those of the authors and do not necessarily represent the official position of the US National Institutes of Health.

## Author Contributions

H.Yu and K.Sun conceived, designed, and supervised the study. X.Deng, Y.Tian, and J.Zou collected the data. X.Deng and K.Sun contributed to the methodology, designed the model. X.Deng and Y.Tian analyzed the model output. X.Deng, Y.Tian, K.Sun, and H.Yu interpreted the results. X.Deng and Y.Tian prepared the figures and tables. X.Deng and Y.Tian wrote the first draft of the manuscript. K.Sun, J.Yang and H.Yu critically revised the content. All authors approved the final manuscript as submitted and agree to be accountable for all aspects of the work.

## Funding

This work was supported by the Key Program of the National Natural Science Foundation of China under Grant 82130093.

## Conflicts of Interest

H.Yu received research funding from Sanofi Pasteur, GlaxoSmithKline, Yichang HEC Changjiang Pharmaceutical Company, Shanghai Roche Pharmaceutical Company, and SINOVAC Biotech Ltd. None of these are related to this study.

## Supplementary Materials

Fig. S1. Distribution of the interval from onset to report. (a) The probability density function. (b) The cumulative distribution function.

Fig. S2. Validation of the importing risk estimation for 24 countries reporting importation cases more than three times.

Fig. S3. Distribution of the number of sexual partners within 1 year among MSM community in mainland China.

Fig. S4. Correlation between the confirmed mpox cases number and active MSM population size of 31 provinces in mainland China.

Table S1. Summary of epidemiological and clinical features.

Table S2. Summary of case number and population of each metropolitan areas among top 12 countries affected by mpox.

Table S3. Summary of international airports for each metropolitan area. Table S4. Fitting information for duration of rash.

Table S5. Case Definitions of mpox in mainland China.

Table S6. Summary of diagnostic accuracy of confirmation test. Table S7. R^2^ for different regression combination.

Table S8. Country/region ISO Alpha-3 code and the according UN name.

## Notes

### Summary of Updates

We update the results of correlation between the confirmed mpox cases number and active MSM population size, and regression models for mpox cases, active MSM population size and air-travel volume according to the updated confirmed mpox cases number from June to September, 2023.

