## Supplemental information for "The Risk of Mpox (Monkeypox) Importation and Subsequent Outbreak Potential in Mainland China: A Retrospective Statistical Modelling Study"

**Abstract**

The 2022 mpox outbreak has spread rapidly across multiple countries in the non-endemic region, mainly among men who have sex with men (MSM), while China only has limited recorded importation and no local outbreak. We constructed probabilistic models to simulate the risk of mpox importation in mainland China, with the help of reported monkeypox cases during this multi-country outbreak and the international air-travel data. And we further evaluated the mpox outbreak potential given that undetected mpox infections were introduced into men who have sex with men, considering different transmissibility, population immunity and population activity. We found that the reduced international air-travel volume and stringent border entry policy decreased about 94% and 69% mpox importations respectively. Once a mpox case is introduced into active MSM population with almost no population immunity, the risk of triggering local transmission is estimated at 42%, and would rise to >95% with over six cases. Our study demonstrates the key role of the reduced international air-travel volume and stringent border entry policy during the COVID-19 pandemic on reducing mpox importations, and the subsequent risk of triggering local outbreaks among MSM.

### 1. Epidemiological and clinical features of mpox

We summarized 12 published literatures, including 3 from endemic countries (2 from Democratic Republic of the Congo, DRC; 1 from Nigeria), 1 from 2003 USA imported outbreak, and 8 from the recent outbreak in Portugal, Spain, UK, etc. And we searched one relavant literature(*1*) review, which show similar result with us. The detailed information can be seen in Table S1.

Table S1. Summary of epidemiological and clinical features.

| **Literature** | **No. of cases** | **Location** | **Period** | **Confirmation** | **Sex** | | **HIV infection** | **Symptoms** | | |
| --- | --- | --- | --- | --- | --- | --- | --- | --- | --- | --- |
|  |  |  |  |  | **Male** | **Female** |  | **Fever** | **Rash** | **Genital rash** |
| Osadebe et al.(*2*) | 333 | Tshuapa Province, DRC ^a^ | Sept 2009-Feb 2014 | PCR | 178/333 (53.4%) ^b^ | 154/333 (44.3%) ^b^ | - | 329/329 (100.0%) | 316/332 (95.2%) | 87/309 (28.2%) |
| Whitehouse et al.(*3*) | 1057 | Tshuapa Province, DRC ^a^ | 2011-2015 | PCR | 568/1054 (53.9%) | 486/1054 (46.1%) | - | 1023/1029 (99.4%) | 1057/1057 (100.0%) | 300/1057 (28.4%) |
| Yinka-Ogunleye et al.(*4*) | 122 ^c^ | Nigeria | Sept 22, 2017-Sept 16, 2018 | PCR/Antibody detection/Viral isolation | 84/122 (68.9%) | 38/122 (31.1%) | 4 ^d^ | 122/122 (100.0%) | 81/92 (88.4%) | 44/65 (67.7%) |
| Huhn et al.(*5*) | 34 | USA | 2003 | PCR/ Viral isolation /Electron microscopy/Antibody detection | 18/34 (52.9%) | 16/34 (47.1%) | - | 29/34 (85.3%) | 33/34 (97.1%) | - |
| Perez Duque et al.(*6*) | 27 | Portugal | May 1-23, 2022 | PCR | 27/27 (100.0%) | 0/27 (0.0%) | 14/26 (53.8%) | 13/26 (50.0%) | 14/15 (93.3%) | 6/15 (40.0%) |
| Inigo Martinez et al.(*7*) | 508 | Madrid, Spain | May 17-Jun 22, 2022 | PCR | 503/508 (99.0%) | 5/508 (1.0%) | 225/508 (44.3%) | 324/508 (63.8%) | 498/508 (98.0%) | 359/508 (72.1%) |
| Patel et al.(*8*) | 197 | London, UK | May 13-July 1, 2022 | PCR | 197/197 (100.0%) | 0/197 (0.0%) | 70/197 (35.5%) | 122/197 (61.9%) | 197/197 (100.0%) | 111/197 (56.3%) |
| Thornhill et al.(*9*) | 528 | 43 sites in 16 countries | Apr 27-Jun 24, 2022 | PCR | 527/528 (99.8%) ^e^ | 0/528 (0.0%) ^e^ | 218/528 (41.3%) | 330/528 (62.5%) | 500/528 (94.7%) | 383/528 (72.5%) ^f^ |
| Philpott et al.(*9*) | 1195 | 43 states, Puerto Rico and DC, USA | May 17-July 22, 2022 | PCR/Next-Generation sequencing/Viral isolation | 1178/1195 (98.7%) ^g^ | 5/1195 (0.4%) ^g^ | 136/334 (41.0%) | 596/941 (63.3%) | 1004/1004 (100.0%) | 333/718 (46.4%) |
| Tarín-Vicente et al.(*10*) | 181 | Madrid and Barcelona, Spain | May 11-Jun 29, 2022 | PCR | 175/181 (97.0%) | 6/181 (3.0%) | 72/181 (40.0%) | 131/181 (72.0%) | 181/181 (100.0%) | 100/181 (55.0%) |
| Català et al.(*11*) | 185 | Spain | May 28-July 14, 2022 | PCR | 185/185 (100.0%) | 0/185 (0.0%) | 78/185 (42.0%) | 100/185 (54%) | 185/185 (100.0%) | 98/185 (53.0%) |
| Hoffmann et al.(*12*) | 546 | Germany | May 19-Jun 30, 2022 | PCR | 546/546 (100.0%) | 0/546 (0.0%) | 256/546 (46.9%) | 272/511 (53.2%) | 493/546 (90.3%) | 267/535 (49.9%) |

a. Democratic Republic of the Congo.
b. One missing.
c. Both confirmed and probable case (any suspected case in whom laboratory testing could not be done but who could be epidemiologically linked with a confirmed case).
d. Four of the people who died had HIV with features of AIDS.
e. One nonbinary/trans.
f. Anogenital area.
g. Three transgender men, five transgender women and 4 prefer not to answer.

### 2. Metropolitan areas and international airports chosen for analysis from top 12 countries

As described in the main text, we assume the travel-associated mpox cases that contribute to the international spread mostly reside in major metropolitan areas with international airport transportation hubs. Consequently, we chose the top 12 countries affected by the 2022 mpox outbreak which account for 90.3% of all mpox cases (as of September 11, 2022). For each country, we focused on the metropolitan areas with most confirmed cases along with major international airports. A full list of selected metropolitan areas, the reported number of mpox cases, along with their corresponding international airports were repored in Table S2 and S3.

Table S2. Summary of case number and population of each metropolitan areas among top 12 countries affected by mpox.

| **Country** | **Date of cases reported** | **Cumulative cases (N)** | **Metropolitan** | **Cases within metropolitan (n)** | **Percent (n/N*100%)** | **Population** |
| --- | --- | --- | --- | --- | --- | --- |
| Total | - | 58961 | - | 39114 | 66.34 | 268066081 |
| Brazil ^a^ | 2022/9/13 | 5559(*13*) | Goiânia | 238 | 4.28 | 1555626(*14*) |
|  |  |  | São Paulo | 2301 | 41.39 | 12396372 |
|  |  |  | Rio de Janeiro | 570 | 10.25 | 6775561 |
| Canada ^a^ | 2022/10/7 | 1411(*15*) | Montreal | 449 ^b^ | 31.82 | 4291732(*16*) |
|  |  |  | Ottawa–Gatineau | 41 | 2.91 | 1488307 |
|  |  |  | Toronto | 496 | 35.15 | 6202225 |
|  |  |  | Vancouver | 165 | 11.69 | 2642825 |
| Colombia ^a^ | 2022/10/10 | 1653(*17*) | Antioquia | 288 | 17.42 | 5974788(*18*) |
|  |  |  | Bogota | 1148 | 69.45 | 7181469 |
| France ^a^ | 2022/9/12 | 3898(*19*) | Ile-de-France | 2332 | 61.11 | 12400000(*20*) |
| Germany ^a^ | 2022/10/13 | 3653(*21*) | Berlin | 1660 | 45.44 | 3677472(*22*) |
|  |  |  | North Rhine-Westphalia | 796 | 21.79 | 17924591 |
| Italy ^a^ | 2022/10/11 | 856(*23*) | Lazio | 151 | 17.64 | 5715190(*24*) |
|  |  |  | Lombardia | 350 | 40.89 | 9965046 |
| Netherlands ^a^ | 2022/10/7 | 1219(*25*) | Noord-Holland & Flevoland | 713 | 58.49 | 3316712(*26*) |
| Peru ^a^ | 2022/9/17 | 2091(*27*) | Lima Metropolitan | 1648 | 78.81 | 10883000(*28*) |
| Portugal ^a^ | 2022/9/14 | 839(*29*) | Lisboa e Vale do Tejo | 650 | 77.47 | 2869627(*30*) |
| Spain ^a^ | 2022/9/16 | 7239(*31*) | Catalonia | 2149 | 29.69 | 7679410(*32*) |
|  |  |  | Madrid | 2500 | 34.54 | 6769113 |
| United Kingdom ^a^ | 2022/9/13 | 3552(*33*) | London | 2335 | 65.74 | 8800000(*34*) |
| United States ^c^ | 2022/10/7 | 26991(*35*) | Atlanta-Sandy Springs-Alpharetta, GA MSA | 1437 ^d^ | 5.32 | 6087762(*36*) |
|  |  |  | Boston-Cambridge-Newton, MA-NH MSA | 373 ^d^ | 1.38 | 4878211 |
|  |  |  | Chicago-Naperville-Elgin, IL-IN-WI MSA | 257 | 0.95 | 9515232 |
|  |  |  | Dallas-Fort Worth-Arlington, TX MSA | 989 ^e^ | 3.66 | 7694138 |
|  |  |  | Denver-Aurora-Lakewood, CO MSA | 221 ^d^ | 0.82 | 2991231 |
|  |  |  | Detroit–Warren–Dearborn, MI MSA | 124 | 0.46 | 4304136 |
|  |  |  | Houston-The Woodlands-Sugar Land, TX MSA | 928 ^e^ | 3.44 | 7154478 |
|  |  |  | Los Angeles-Long Beach, CA CSA | 2607 | 9.66 | 17788274 |
|  |  |  | Miami-Fort Lauderdale-Pompano Beach, FL MSA | 1112 | 4.12 | 6173008 |
|  |  |  | Minneapolis-St. Paul-Bloomington, MN-WI MSA | 207 ^g^ | 0.77 | 3657477 |
|  |  |  | New York-Newark-Jersey City, NY-NJ-PA MSA | 4381 ^f, g^ | 16.23 | 19068287 |
|  |  |  | Philadelphia-Camden-Wilmington, PA-NJ-DE-MD MSA | 774 ^d, g^ | 2.87 | 6107906 |
|  |  |  | Phoenix-Mesa-Chandler, AZ MSA | 398 | 1.47 | 5059909 |
|  |  |  | Sacramento-Roseville-Folsom, CA MSA | 178 ^g^ | 0.66 | 2650900 |
|  |  |  | San Diego-Chula Vista-Carlsbad, CA MSA | 404 | 1.50 | 3332427 |
|  |  |  | San Jose-San Francisco-Oakland, CA CSA | 1400 ^g^ | 5.19 | 6668062 |
|  |  |  | Seattle-Tacoma-Bellevue, WA MSA | 550 | 2.04 | 4018598 |
|  |  |  | Tampa-St. Petersburg-Clearwater, FL MSA | 213 | 0.79 | 3243963 |
|  |  |  | Washington-Baltimore-Arlington, DC-MD-VA-WV-PA CSA | 1581 ^d^ | 5.85 | 9163016 |

a. The case number of the metropolitan area was approximately regarded as that reported to the major city (sharing the same name with the metropolitan area).
b. Due to no report at a metro-level in Qubec, Canada, the case number of Montreal was estimated by multiplying the total number of mpox cases in Quebec by the proportion of Toronto’s cases in Ontario.
c. The case number of a metropolitan statistical area (MSA) or combined statistical areas (CSA) is the sum of all counties’ cases within the MSA/CSA.
d. For these metropolitan areas, case number were not available at county level, but only available at the state level. We assume that the proportion of mpox cases in the metropolitan areas with repect to the state was the same as the proportion of MSM population in the metropolitan area with respect to the state’s MSM population. The county-level MSM population estimates for each county were based on Grey et al(*37*).
e. For Texas, official report was at a level of Public Health Region (PHR). The cases were estimated by multiplying the ratio of MSM of the county to that of the PHR with the PHR’s cases.
f. Pike County was unavailable and removed.
g. Reported case number of <5 or <11 was regarded as 5 or 11, respectively.

Table S3. Summary of international airports for each metropolitan area.

| **Country** | **Metropolitan** | **Airport** | **OAG code** |
| --- | --- | --- | --- |
| Brazil | Goiânia | Goiânia International Airport | GYN |
|  | Rio de Janeiro | Rio de Janeiro/Galeão International Airport | GIG |
|  |  | Jacarepaguá Airport | RRJ |
|  |  | Santos Dumont Airport | SDU |
|  | São Paulo | São Paulo–Congonhas Airport | CGH |
|  |  | São Paulo/Guarulhos International Airport | GRU |
|  |  | Jundiaí Airport | QDV |
|  |  | São José dos Campos Airport | SJK |
|  |  | Viracopos International Airport | VCP |
| Canada | Montreal | Montréal–Mirabel International Airport | YMX |
|  |  | Montreal Saint-Hubert Longueuil Airport | YHU |
|  |  | Montréal–Trudeau International Airport | YUL |
|  | Ottawa–Gatineau | Gatineau-Ottawa Executive Airport | YND |
|  |  | Ottawa Macdonald–Cartier International Airport | YOW |
|  | Toronto | Billy Bishop Toronto City Airport | YTZ |
|  |  | Buttonville Municipal Airport | YKZ |
|  |  | John C. Munro Hamilton International Airport | YHM |
|  |  | Region of Waterloo International Airport | YKF |
|  |  | Toronto Pearson International Airport | YYZ |
|  | Vancouver | Abbotsford International Airport | YXX |
|  |  | Boundary Bay Airport | YDT |
|  |  | Vancouver International Airport | YVR |
| Colombia | Antioquia | José María Córdova International Airport | MDE |
|  |  | Olaya Herrera Airport | EOH |
|  | Bogota | Bogota FAirport | BOG |
| France | Ile-de-France | Beauvais–Tillé Airport | BVA |
|  |  | Châlons Vatry Airport | XCR |
|  |  | Charles de Gaulle Airport | CDG |
|  |  | Le Bourget Airport | LBG |
|  |  | Orly Airport | ORY |
| Germany | Berlin | Brandenburg Airport | BER |
|  |  | Schönefeld Airport | SXF |
|  |  | Tegel Airport | TXL |
|  | North Rhine-Westphalia | Cologne Bonn Airport | CGN |
|  |  | Dortmund Airport | DTM |
|  |  | Düsseldorf Airport | DUS |
|  |  | Münster Osnabrück International Airport | FMO |
|  |  | Weeze Airport | NRN |
| Italy | Lazio | Ciampino–G. B. Pastine International Airport | CIA |
|  |  | Leonardo da Vinci–Fiumicino Airpor | FCO |
|  | Lombardia | Linate Airport | LIN |
|  |  | Malpensa Airport | MXP |
|  |  | Orio al Serio International Airport | BGY |
|  |  | Parma Airport | PMF |
| Netherlands | Noord-Holland & Flevoland | Amsterdam Schiphol airport | AMS |
| Peru | Lima | Jorge Chávez International Airport | LIM |
| Portugal | Lisboa e Vale do Tejo | Lisbon Airport | LIS |
| Spain | Madrid | Adolfo Suárez Madrid–Barajas Airport | MAD |
|  | Catalonia | Josep Tarradellas Barcelona–El Prat Airport | BCN |
|  |  | Girona–Costa Brava Airport | GRO |
|  |  | Lleida–Alguaire Airport | ILD |
|  |  | Reus Airport | REU |
| United Kingdom | London | Gatwick Airport | LGW |
|  |  | Heathrow Airport | LHR |
|  |  | London City Airport | LCY |
|  |  | London Southend Airport | SEN |
|  |  | London Stansted Airport | STN |
|  |  | Luton Airport | LTN |
| United States | Atlanta-Sandy Springs-Alpharetta, GA MSA | DeKalb–Peachtree Airport | PDK |
|  |  | Fulton County Airport (Georgia) | FTY |
|  |  | Gwinnett County Airport | LZU |
|  |  | Hartsfield–Jackson Atlanta International Airport | ATL |
|  | Boston-Cambridge-Newton, MA-NH MSA | Beverly Regional Airport | BVY |
|  |  | Cambridge–Dorchester Airport | CGE |
|  |  | Hanscom Field | BED |
|  |  | Lawrence Municipal Airport (Massachusetts) | LWM |
|  |  | Logan International Airport | BOS |
|  |  | Manchester–Boston Regional Airport | MHT |
|  |  | Newton City/County Airport | EWK |
|  |  | Norwood Memorial Airport | OWD |
|  |  | Rhode Island T. F. Green International Airport | PVD |
|  |  | Worcester Regional Airport | ORH |
|  | Chicago-Naperville-Elgin, IL-IN-WI MSA | Chicago Executive Airport | PWK |
|  |  | Chicago Rockford International Airport | RFD |
|  |  | DuPage Airport | DPA |
|  |  | Midway International Airport | MDW |
|  |  | Milwaukee Mitchell International Airport | MKE |
|  |  | O'Hare International Airport | ORD |
|  | Dallas-Fort Worth-Arlington, TX MSA | Addison Airport | ADS |
|  |  | Dallas Fort Worth International Airport | DFW |
|  |  | Dallas Love Field | DAL |
|  |  | Fort Worth Alliance Airport | AFW |
|  |  | Fort Worth Meacham International Airport | FTW |
|  | Denver-Aurora-Lakewood, CO MSA | Centennial Airport | APA |
|  |  | Aurora Municipal Airport | AUZ |
|  |  | Rocky Mountain Metropolitan Airport | BJC |
|  |  | Denver International Airport | DEN |
|  | Detroit–Warren–Dearborn, MI MSA | Ann Arbor Municipal Airport | ARB |
|  |  | Coleman A. Young International Airport | DET |
|  |  | Detroit Metropolitan Airport | DTW |
|  |  | Bishop International Airport | FNT |
|  |  | St. Clair County International Airport | PHN |
|  |  | Oakland County International Airport | PTK |
|  |  | Willow Run Airport | YIP |
|  | Houston-The Woodlands-Sugar Land, TX MSA | David Wayne Hooks Memorial Airport | DWH |
|  |  | Ellington Airport (Texas) | EFD |
|  |  | William P. Hobby Airport | HOU |
|  |  | George Bush Intercontinental Airport | IAH |
|  |  | West Houston Airport | IWS |
|  |  | Sugar Land Regional Airport | SGR |
|  | Los Angeles-Long Beach, CA CSA | Hollywood Burbank Airport | BUR |
|  |  | Los Angeles International Airport | LAX |
|  |  | Long Beach Airport | LGB |
|  |  | Ontario International Airport | ONT |
|  |  | San Bernardino International Airport | SBD |
|  |  | John Wayne Airport | SNA |
|  |  | Van Nuys Airport | VNY |
|  | Miami-Fort Lauderdale-Pompano Beach, FL MSA | Boca Raton Airport | BCT |
|  |  | Fort Lauderdale–Hollywood International Airport | FLL |
|  |  | Fort Lauderdale Executive Airport | FXE |
|  |  | Homestead Air Reserve Base | HST |
|  |  | Miami International Airport | MIA |
|  |  | Miami-Opa Locka Executive Airport | OPF |
|  |  | Palm Beach International Airport | PBI |
|  |  | Pompano Beach Airpark | PPM |
|  |  | Miami Executive Airport | TMB |
|  |  | Dade-Collier Training and Transition Airport | TNT |
|  | Minneapolis-St. Paul-Bloomington, MN-WI MSA | Flying Cloud Airport | FCM |
|  |  | Crystal Airport (Minnesota) | MIC |
|  |  | Minneapolis–Saint Paul International Airport | MSP |
|  |  | St. Paul Downtown Airport | STP |
|  | New York-Newark-Jersey City, NY-NJ-PA MSA | Newark Liberty International Airport^a^ | EWR |
|  |  | Westchester County Airport | HPN |
|  |  | Tweed New Haven Airport | HVN |
|  |  | Long Island MacArthur Airport | ISP |
|  |  | John F. Kennedy International Airport | JFK |
|  |  | LaGuardia Airport | LGA |
|  |  | New York Skyports Seaplane Base | NYS |
|  |  | Stewart International Airport | SWF |
|  |  | Trenton–Mercer Airport^a^ | TTN |
|  | Philadelphia-Camden-Wilmington, PA-NJ-DE-MD MSA | Lehigh Valley International Airport | ABE |
|  |  | Atlantic City International Airport | ACY |
|  |  | Wilmington Airport (Delaware) | ILG |
|  |  | Philadelphia International Airport | PHL |
|  |  | Northeast Philadelphia Airport | PNE |
|  |  | Reading Regional Airport | RDG |
|  | Phoenix-Mesa-Chandler, AZ MSA | Phoenix–Mesa Gateway Airport | AZA |
|  |  | Phoenix Deer Valley Airport | DVT |
|  |  | Phoenix Goodyear Airport | GYR |
|  |  | Falcon Field (Arizona) | MSC |
|  |  | Phoenix Sky Harbor International Airport | PHX |
|  |  | Scottsdale Airport | SCF |
|  | Sacramento-Roseville-Folsom, CA MSA | Sacramento McClellan Airport | MCC |
|  |  | Sacramento Mather Airport | MHR |
|  |  | Sacramento Executive Airport | SAC |
|  |  | Stockton Metropolitan Airport | SCK |
|  |  | Sacramento International Airport | SMF |
|  | San Diego-Chula Vista-Carlsbad, CA MSA | McClellan–Palomar Airport | CLD |
|  |  | Montgomery-Gibbs Executive Airport | MYF |
|  |  | San Diego International Airport | SAN |
|  |  | Brown Field Municipal Airport | SDM |
|  |  | Gillespie Field | SEE |
|  | San Jose-San Francisco-Oakland, CA CSA | Oakland International Airport | OAK |
|  |  | Reid–Hillview Airport | RHV |
|  |  | San Francisco International Airport | SFO |
|  |  | San Jose International Airport | SJC |
|  | Seattle-Tacoma-Bellevue, WA MSA | Boeing Field | BFI |
|  |  | Kenmore Air Harbor Seaplane Base | LKE |
|  |  | Paine Field | PAE |
|  |  | Renton Municipal Airport | RNT |
|  |  | Seattle–Tacoma International Airport | SEA |
|  |  | Tacoma Narrows Airport | TIW |
|  | Tampa-St. Petersburg-Clearwater, FL MSA | Clearwater Air Park Airport | CLW |
|  |  | St. Pete–Clearwater International Airport | PIE |
|  |  | Albert Whitted Airport | SPG |
|  |  | Tampa International Airport | TPA |
|  |  | Peter O. Knight Airport | TPF |
|  | Washington-Baltimore-Arlington, DC-MD-VA-WV-PA CSA | Baltimore/Washington International Airport | BWI |
|  |  | Ronald Reagan Washington National Airport | DCA |
|  |  | Washington Dulles International Airport | IAD |
|  |  | Martin State Airport | MTN |

a. These airports serve both New York-Newark-Jersey City, NY-NJ-PA MSA and Philadelphia-Camden-Wilmington, PA-NJ-DE-MD MSA. Here for simplicity, we assume these airports only serve NY-NJ-PA MSA according to their locations.

### 3. Mpox incubation period, duration of rash presentation, and onset to reporting delays

Based on the estimation by Miura et al(*38*), the incubation period of mpox infection in the Netherlands had a median of 8.5 days with 5^th^–95^th^ percentiles of 4.2–17.3 days. The distribution of the incubation period is best fitted by a lognormal distribution. We assume the duration from infection to symptom onset follows the same distribution.

The duration of rash is estimated using the empirical data from Huhn et al(*5*). We assume the range as the 2.5^th^ and 97.5^th^ percentiles. Lognormal, Gamma, and Weibull distribution are fitted using nonlinear least squares method. The distribution with the minimum sum of square residual is chosen as the distribution for the duration of rash (Table S4).

Table S4. Fitting information for duration of rash.

| **Distribution** | **Parameter 1** | **Parameter 2** | **Residual (sum of square)** |
| --- | --- | --- | --- |
| Gamma | 15.32373478 | 1.248809497 | 0.000572 |
| Lognormal | 2.486530301 | 0.29177409 | 0.000313 |
| Weibull | 6.140548705 | 12.73805504 | 0.000625 |

### 4. Mpox onset to reporting delays

We downloaded the empirical data of the time interval from onset to reporting from the WHO situation dashboard(*39*). Then, we fit the data using zero-inflated passion (hereafter ZIP for short) and zero-inflated negative binomial (hereafter ZINBI for short) through a maximum likelihood method. The fitting of ZINBI is better, which can be seen in Fig. S1.

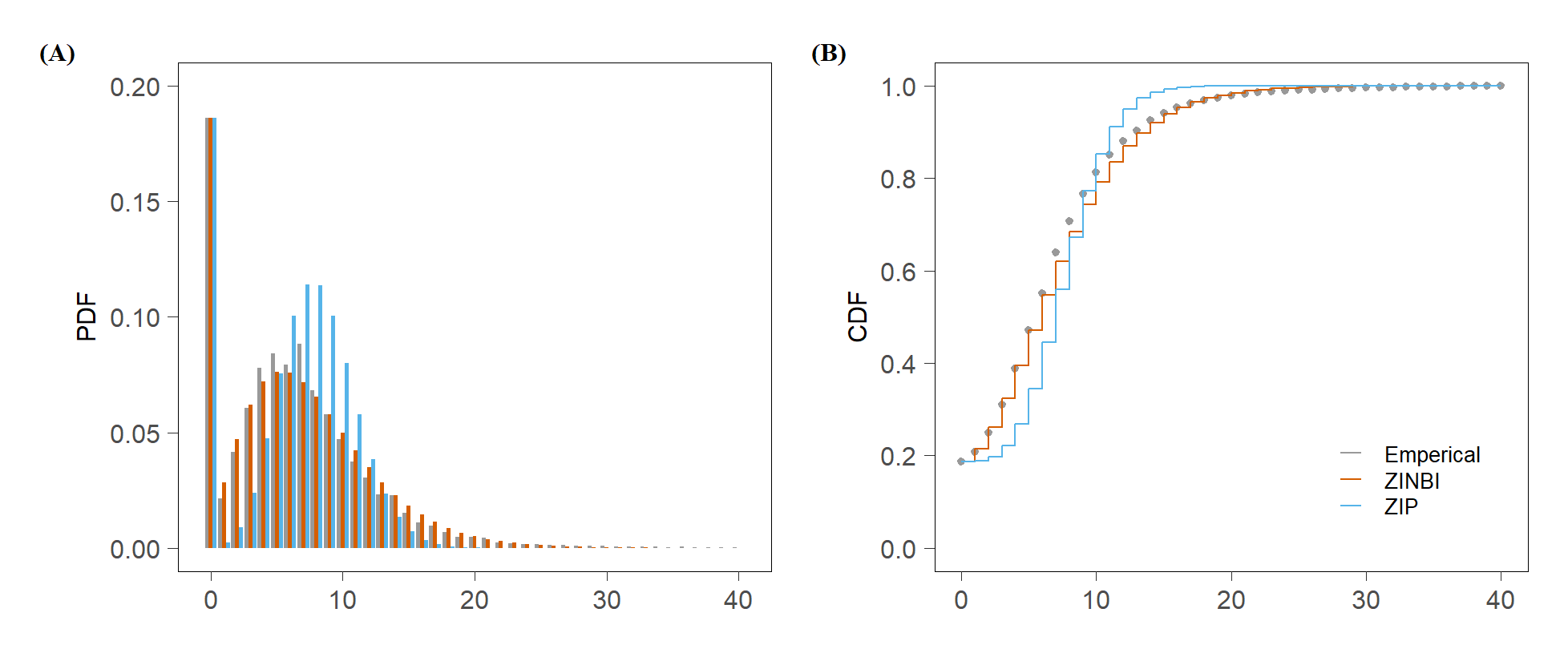

Fig. S1. Distribution of the interval from onset to report. (a) The probability density function. (b) The cumulative distribution function.

### 5. Validation of the importation risk estimation

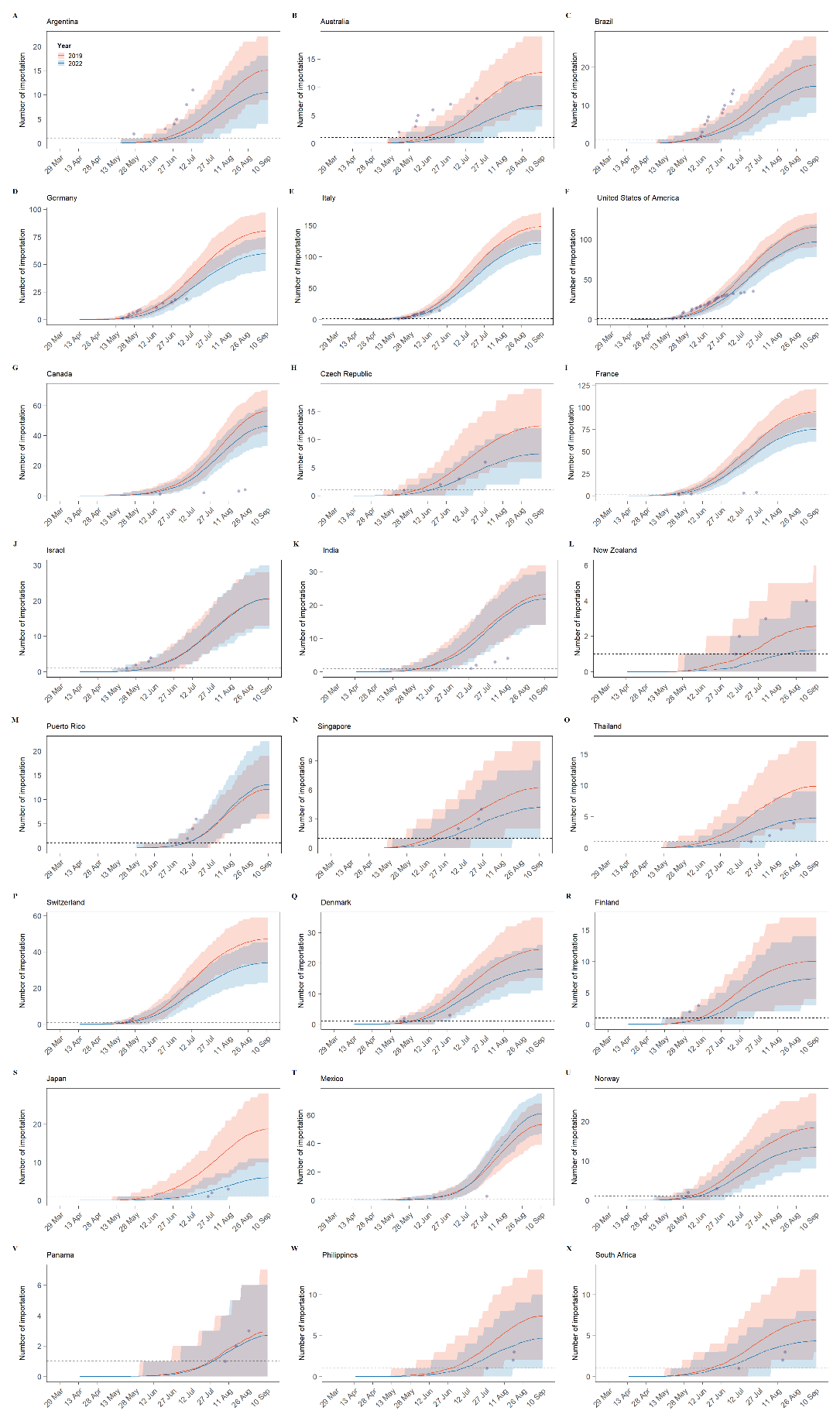

Fig. S2. Validation of the importing risk estimation for 24 countries reporting importation cases more than three times. In each panel, the red ribbon indicates the estimation based on air-travel volumn of 2019 (upper bound estimation) and the blue one indicates that of 2022 (lower bound estimation). Purple dots indicate the observed importations.

### 6. Mpox Case Definitions

Table S5. Case Definitions of mpox in mainland China.

| Class | Definitions |
| --- | --- |
| Suspect Case | With symptoms of fever, swollen lymph nodes and skin or mucosal rash and with any of the following epidemiological history:  Within 21 days of illness onset:   - Having overseas travel history in areas with mpox case reports. - Having close contact with a confirmed case of mpox. - Exposed to blood, body fluids and secretions of mpox-infected animals such as rodents and non-human primates. |
| Confirmed Case | Meeting the definition of suspected cases, laboratory tests, mpox virus nucleic acid positive or virus isolation positive |

### 7. Border screening

In Scenario 1 corresponding to the mainland China’s quarantine policy till June 28, 2022, we consider all international travelers arriving at mainland China need to undergo an entry quarantine, but with varying quarantine duration through the year of 2022 due to policy adjustment over time(*40*). In Scenario 2, only individuals with known exposure to mpox need to undergo medical observations for mpox related symptoms. During the medical observation, exposed individuals need to abstain from sexual activity, along with other precautious measures. No mandatory isolation is required unless exposed individuals start developing mpox related symptoms during the period of medical observation. Both confirmed and suspected mpox cases need to be isolated at the designated medical facilities and we assume they could not further transmit mpox to others in our analysis.

We simulated 100 mpox infected entry personals, among whom 90% would develop rash following a Bernoulli process. We assigned each individual’s the incubation period according to a lognormal distribution(*38*) with mean of 6.48 days and the standard deviation of 28.2 days. We assume the duration of rash following a lognormal (Table S4) with mean of 2.49 days and the standard deviation of 0.29 days. Then, we randomly assigned the border entry date of each infected indivudal with unifom likelihood within the time window of his/her incubation. For Scenario 1, where a 7-day mandatory quarantine is required, mpox PCR test would be performed once rash onset. On the contrary, for Scenario 2, only people reporting the epidemiological link would seek tests once rash onset. For each scenario, we simulated the performance of PCR test following a Bernoulli process with 95% sensitivity based on three published articles(*41-43*). We repeated the entire simulation for 200 times to obtain the distribution.

The probability of mpox infection detection was presented as a function of the COVID-19 quarantine duration and the proportion of self-reporting epidemiological links for Scenario 1. As for Scenario 2, the effectiveness of the border screening policy was explored as a function of the proportion of self-reporting epidemiological links and the duration of the medical observation among imported mpox infections.

The sensitivity of PCR confirmation of mpox is based on the summary of three articles described in Table S6.

Table S6. Summary of diagnostic accuracy of confirmation test.

| **Ref** | **Davi et al.(*41*)** | | **Li et al. 2010.(*42*)** | | | **Li et al. 2006.(*43*)** |
| --- | --- | --- | --- | --- | --- | --- |
| Method | RPA^a^ | rt-PCR | rt-PCR | | | rt-PCR |
| Target | G2R | G2R-G | G2R-WA | G2R-G | C3L | B6R |
| Purpose | both clades of MPXV | | West African clade | both clades of MPXV | Congo Basin clade | MPXV |
| Specimen | Whole blood/serum | | liver and skin tissues, nasal and oral swabs, lesions and human mpox clinical samples | | | Samples of skin, slides, and swabs. |
| Positive | 25 monkeys and 20 humans | | 4 strains, 10 animals and 3 humans | 11 strains, 20 animals and 7 humans | 7 strains, 10 animals and 4 humans | 1 MPXV DNA samples and 13 human clinical samples |
| Negative | 23 monkeys and 27 humans | | 14 strains, 3 other DNA, 10 animals and 4 humans | 7 strains and 3 other DNA | 11 strains, 3 other DNA, 10 animals and 3 humans | 9 *orthopoxviruses* DNA samples, 15 bacteria DNA samples and 2 VZV^b^ clinical samples |
| Result |  | |  | | |  |
| Sensitivity | 43/45 (95.6%) | 45/45 (100.0%) | 17/17 (100.0%) | 38/38 (100.0%) | 21/21 (100.0%) | 14/14 (100.0%) |
| Specificity | 50/50 (100.0%) | 50/50 (100.0%) | 31/31 (100.0%) | 10/10 (100.0%) | 27/27 (100.0%) | 26/26 (100.0%) |

a. Recombinase polymerase amplification.
b. Varicella zoster virus.

### 8. Outbreak probability

Following Hartfield et al.’s analytical solution of outbreak probability $T_{0}$ of a single seeding case as a function of the basic reproduction number $R_{0}$ and dispersion parameter $k$.

$$T_{0}=\frac{1}{\log R_{0}}(0.334+\frac{0.689}{k}+\frac{0.408}{R_{0}}-\frac{0.507}{kR_{0}}-\frac{0.356}{{R_{0}}^{2}}+\frac{0.467}{k{R_{0}}^{2}})$$

A reasonable guess of the mpox’s basic reproduction number among MSM community in mainland China can be approximated by what’s being observed in other non-endemic countries with substential outbreak size ($R_{0}=1.8$ base a recent study by Kwok et al(*44*)).

We use the distribution of the number of sexual partners among MSM in mainland China to approximate the dispersion of mpox transmission $k$. Fig. S3 displays the observed distribution of the number of sexual partners among MSM in mainland China(*45, 46*). The number of sexual partners ranges from 1 to 53, which can be characterized by a negative binomial distribution with mean *μ* = 3.04 and variance *μ*(1 + *μ*/*k*) = 13.58, where *k* = 0.88 is the dispersion parameter estimated by the distribution fitting.

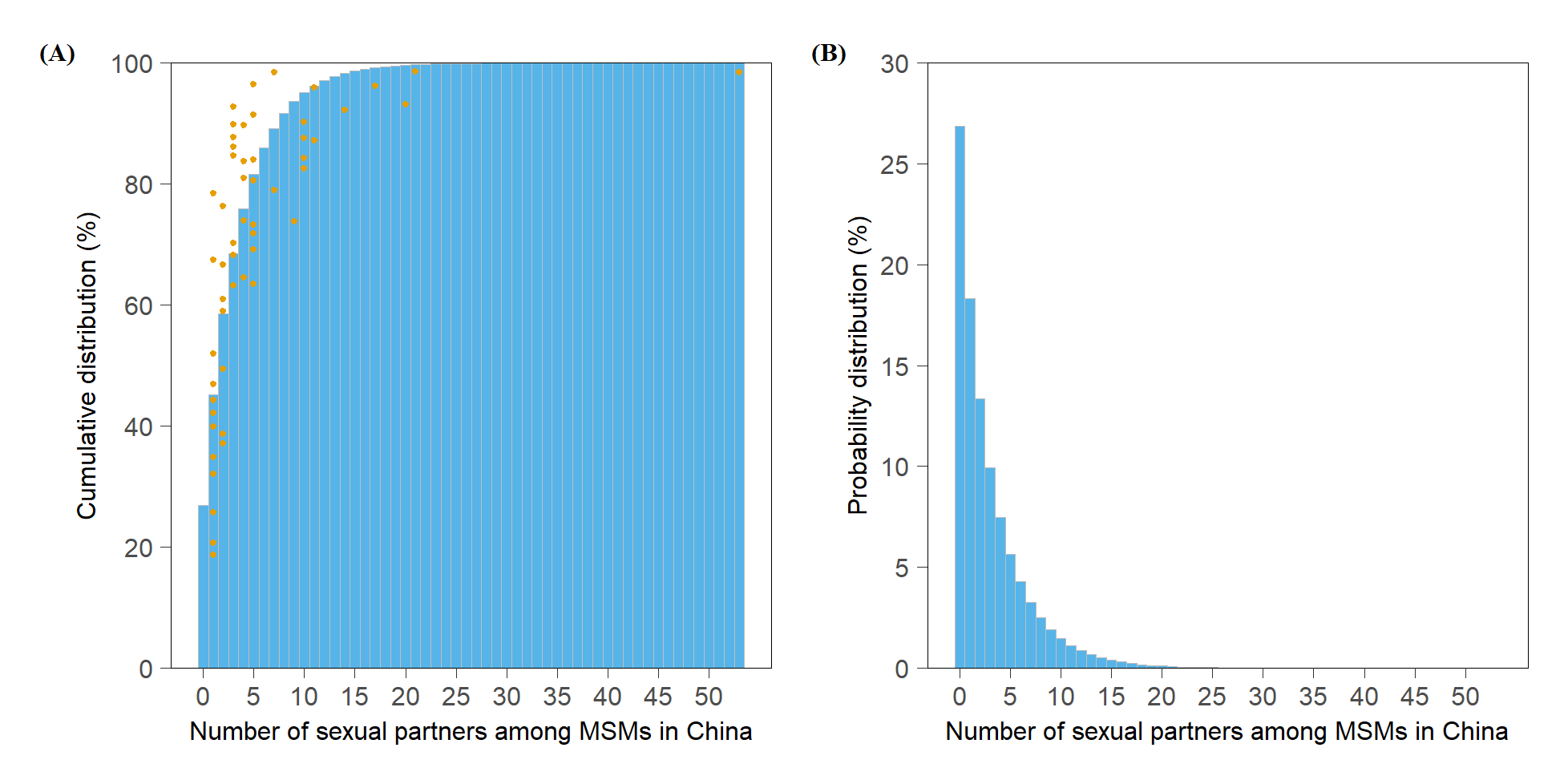

Fig. S3. Distribution of the number of sexual partners within 1 year among MSM community in mainland China. (a) The cumulative distribution in mainland China. (b) The probability mass distribution of the number of MSM sexual partners in mainland China. In panel a, dots in color of yellow indicated raw data extracted from literature(*45, 46*).

### 9. Correlation between the confirmed mpox cases number and active MSM population size

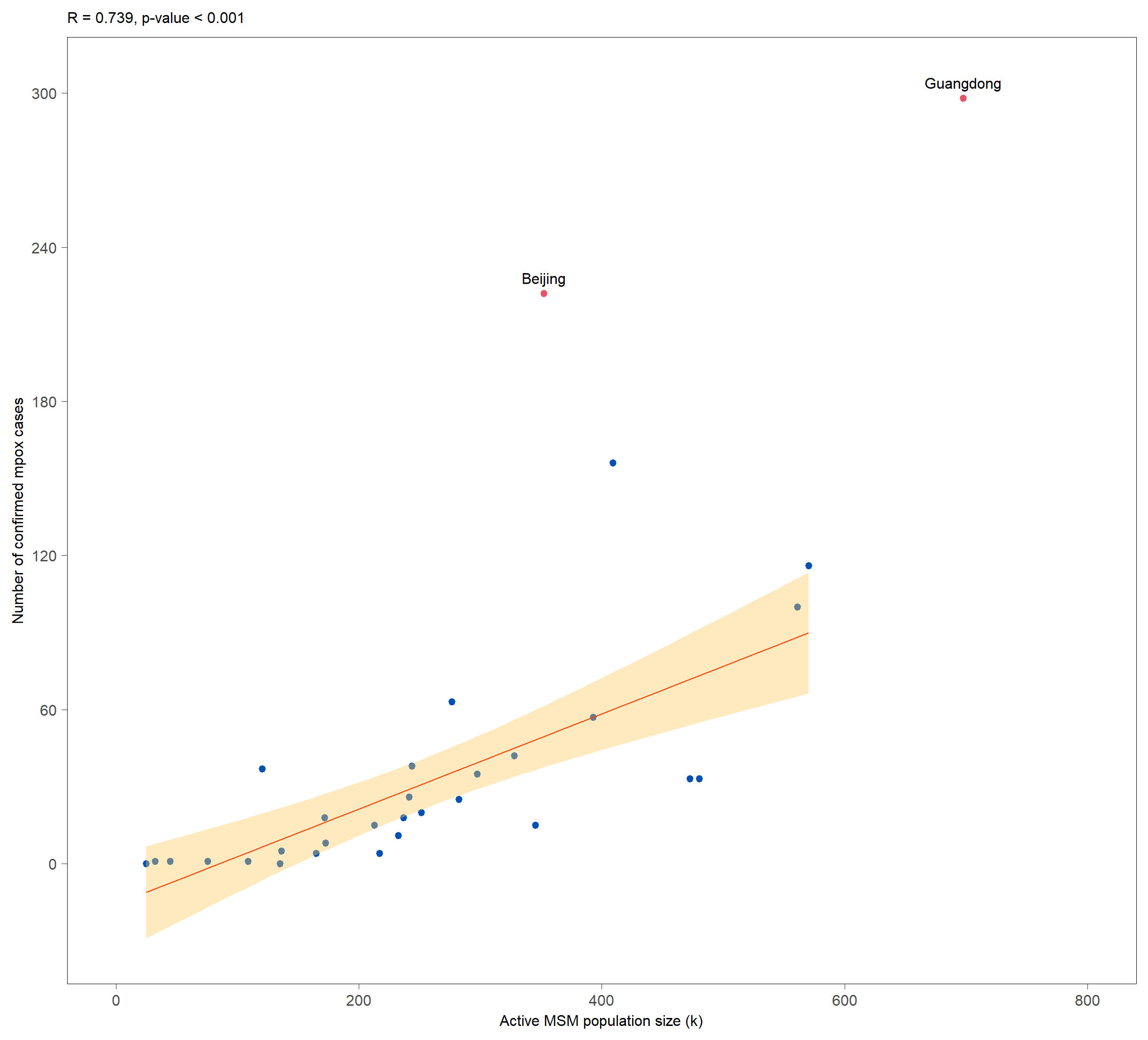

Fig. S4. Correlation between the confirmed mpox cases number and active MSM population size of 31 provinces in mainland China. The orange ribbon indicates the estimation based on the active MSM population size(*47*). The blue dots indicate different provinces in mainland China and the two red dots present outliers in the analysis.

### 10 Regression models for mpox cases, active MSM population size and air-travel volume

Table S7. R^2^ for different regression combination.

| **Var** | **31 provinces** | **29 provinces ^a^** |
| --- | --- | --- |
| active MSM population size | 0.5088 | 0.5288 |
| 2019 air-travel volume | 0.3395 | 0.1294 |
| active MSM population size + 2019 air-travel volume | 0.6223 | 0.5585 |
| active MSM population size + 2019 air-travel volume + active MSM population size * 2019 air-travel volume | 0.8180 | 0.5981 |

a. Excluded Beijing and Guangdong province.

For combination including active MSM population size, 2019 air-travel volume and their interaction term (R^2^ = 0.8452), the formula is described as:

$\mu_{i}=\alpha+\beta_{1}x_{1}+\beta_{2}x_{2}+\beta_{3}x_{1}x_{2}$,

where $\mu_{i}$ is the confirmed mpox cases number, $x_{1}$ is the active MSM population size (100 thousand), $x_{2}$ is the 2019 air-travel volume (100 thousand), $x_{1}x_{2}$ is the interaction term, $\alpha$ is the constant term and $\beta_{i}$ is the coefficient.

### 11. Country/region ISO Alpha-3 code and the according UN name

Table S8. Country/region ISO Alpha-3 code and the according UN name.

| **ISO Alpha-3** | **Name by UN** |
| --- | --- |
| BRA | Brazil |
| CAN | Canada |
| COL | Colombia |
| DEU | Germany |
| ESP | Spain |
| FRA | France |
| GBR | United Kingdom |
| ITA | Italy |
| NLD | Netherlands |
| PER | Peru |
| PRT | Portugal |
| USA | United States of America |
